# What influences COVID-19 infection rates: A statistical approach to identify promising factors applied to infection data from Germany

**DOI:** 10.1101/2020.04.14.20064501

**Authors:** Moritz Mercker, Uwe Betzin, Dennis Wilken

## Abstract

The recent COVID-19 pandemic is of big and world-wide concern. There is an intense discussion and uncertainty which factors and sanctions can reduce infection rates. The overall aim is to prevent an overload of the medical system. Even within one country, there is frequently a strong local variability in both – political sanctions as well as other local factors – which may influence infection rates. The main focus of study is analysis and interpretation of recent temporal developments (infection rates). We present a statistical framework designed to identify local factors which reduce infection rates. The approach is robust with respect to the number of undetected infection cases. We apply the framework to spatio-temporal infection data from Germany. In particular, we demonstrate that (1) infection rates are in average significantly decreasing in Germany; (2) there is a high spatial variability of these rates, and (3) both, early emergence of first infections and high local infection densities has led to strong recent decays in infection rates, suggesting that psychological effects (such as awareness of danger) lead to behaviour changes that reduce infection rates. However, the full potential of the presented method cannot yet be exploited, since more precise spatio-temporal data, such as local cell phone-based mobility data, are not yet available. In the nearest future, the presented framework could be applied to data from other countries at any state of infection, even during the exponential phase of the growth of infection rates.

## Introduction

Since March 2020 the new Disease COVID-19 is declared a pandemic by the World Health Organization (WHO). The disease is caused by a novel virus called severe acute respiratory syndrome (SARS)-CoV-2. Transmission of the virus between humans is known to happen via respiratory droplets and until today, no specific medical treatment for the disease is known. The virus was first reported in Wuhan, Hubei, China, in December 2019. The disease had spread widely over the province of Hubei, but was contained after strict quarantine imposed on January 23, 2020. The peak of daily confirmed cases of about 4000 was reached on February 3, ten days after the quarantine had been imposed; then the number of daily new cases decreased below 100 at the beginning of March [8]. Until today nearly exponential growth has been observed in several countries worldwide including, e.g., Italy, Spain, and the U.S. Despite the exponential increase, several countries have already observed a decrease in the daily number of infections (e.g. South Korea, Italy, Spain, and Germany).

In order to better understand various aspects of the pandemic development, research on COVID-19 exploded during the last months. Besides medical studies, also various statistical and mathematical infection models have been developed and applied to current data, in order to predict possible infection developments under certain conditions/restrictions or to compare infection rates among different countries (see, among others, [1, 4, 10, 11, 13]). However, models often suffer from the fact that certain mechanistic assumptions are critical for the conclusions and predictions, but can often hardly be validated.

In this work, we present a statistical framework which is not based on specific mechanistic infection models, but is constructed to analyse current infection numbers, where the latter may show an arbitrary complex behaviour, possibly varying among different sub-regions of the considered area. The framework is optimised to reveal factors that explain differences in recent infection rates among different sub-regions. Thus, in the light of high variability of possible local political, social, environmental or infrastructural factors that may affect the infection rates, the proposed tool provides an opportunity to extract the most important factors responsible for COVID-19 propagation. This is important not only to reduce the exponential rate of infection growth in different countries, but also to decide which sanctions can be eased if the maximum of the confirmed daily new cases has been surpassed, as it is currently the case in Germany and several other countries.

## Material and Methods

### Data structure and availability

German COVID-19 Infection data have been obtained from the Robert-Koch Institut (RKI) provided via the national platform for geographical data (NPGEO Corona Hub 2020: https://npgeo-corona-npgeo-de.hub.arcgis.com). In particular, data are available in terms of daily reported infections separately for each of the 402 existing administrative districts (ADs), which are nested within 16 federal states (Fig. 1).

**Figure 1.**
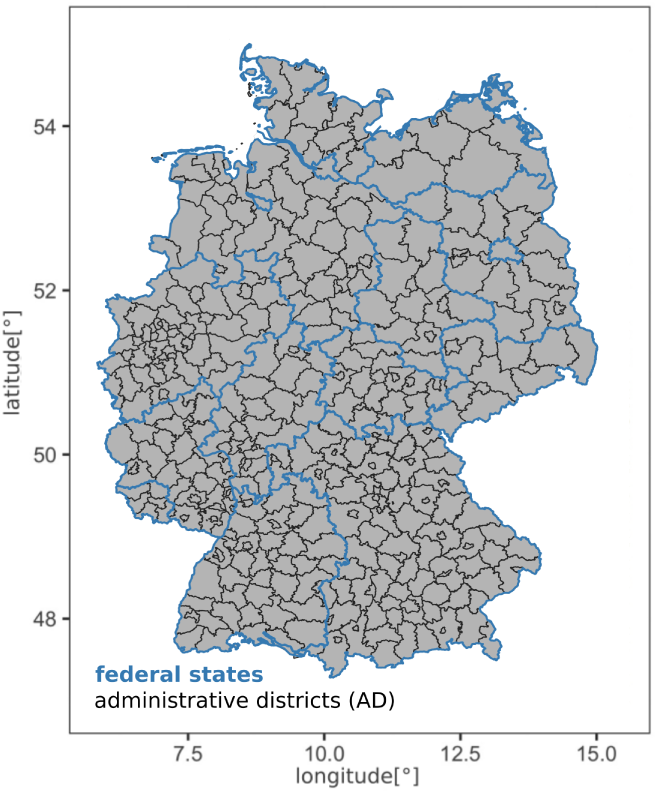
Map of Germany including the frontiers of the federal states (blue lines) and the administrative district (black lines) (data from https://public.opendatasoft.com and https://gdz.bkg.bund.de)

As a start of the time series we have set February the 22th (2020), which was just before the beginning of the pandemic in Germany. The time series considered in this work ends at April 13th (2020).

### Quantifying decrease in infection rates

In order to quantify the AD-related recent change in infection rates, we applied three different partially related methods separately to infection rate time series of each sub-region leading to three different measures (*β*_1_, *β*_2_, and *β*_3_). Their derivation is detailed in the following.

First, we fitted the generalised additive mixed model (GAMM) [7, 21] to the entire time series of each AD, where the federal state has been used as a random intercept, and the date as a smooth predictor term. Additive regression models allow to estimate nonlinear dependencies between predictor variables (here e.g. the date) and an outcome variable (here e.g. the number of reported infections) [7,21]. Consideration of such nonlinearities is fundamental here, since the initial increase and possible later decrease in infection numbers is often strongly nonlinear. In particular (if not stated otherwise) the optimal smoothness has been determined using generalised cross-validation methods [18]. Random intercepts (i.e., the use of mixed modelling [2, 21]) have been applied if data are not independent, but instead nested within certain units (such as states). Since infection numbers are additionally temporally autocorrelated (infection numbers depend to some degree on the number of the day before), an auto-regression (AR) structure of order 1 has been added to the model. Finally, a negative-binomial probability distribution has been used, to account for the fact that infection numbers do not follow a normal distribution but are (possibly overdispersed) count data [9, 20, 21]. In addition to the date, also the weekday has been introduced as a smooth, in particular as a cyclic smooth [18] in order to partial out the distinct effect of the weekday on reported infection numbers.

Second, separately for each AD, the above described GAMM has been used to predict the partial effect of the date, i.e., the (possibly nonlinear) date-dependent development of infection rates, where the effect of the weekday has been eliminated. This smooth predicted curve has been subsequently normalised by dividing all predicted values by the mean. The latter step ensures the comparability of the temporal development of infection numbers between the different ADs.

Finally, the normalised smooth curve has been used for each AD to create two different measures representing the strength of the recent decay: (1) we calculated the value *β*_1_, which is defined by the curve value of the most current date minus the value 10 days before, and (2) we calculated *β*_2_, which is defined by the value of the current data divided by the maximum of the curve. Thus, *β*_1_ is proportional to the current slope of the infection curve (averaged over the last 10 days), whereas *β*_2_ rather relates the current infection number to the overall maximum of the smoothed infection curve. Hence, *β*_1_ is a reasonable measure even if most of the local infection curves are still increasing, whereas *β*_2_ is only appropriate if most of the infection curves are already decreasing.

To calculate a third type of measure for recent changes in infection rates, a second GAMM was fitted to the infection data of each AD – similar to the above described GAMM approach, however with two differences: first, the time series has been restricted to the last 14 days, and second, the dependency on the date has been assumed to be (log-)linear instead of nonlinear. All other model specifications were as described above for the calculation of *β*_1_ and *β*_2_. The advantage of the non-smooth (log-linear) incorporation of the date variable, here, *β*_3_ (the slope of the average log-linear development) was represented by a corresponding date-dependent regression coefficient, and thus confidence intervals respectively standard errors could be computed, the latter representing the certainty of each estimate. Furthermore, after slight reformulations, *β*_3_ can be directly interpreted, namely in terms of average percentage change in infection rates per day. A possible disadvantage of *β*_3_ compared to *β*_1_ and *β*_2_ is that a shorter time series (and thus less data) have been used, which was necessary to select a time frame with nearly log-linear behaviour of infection rates corresponding to the model.

Thus, in summary, three different (partially related) measures have been constructed to characterise the recent infection rate, namely *β*_1_ and *β*_3_ (where a negative value indicates a recent decay) as well as *β*_2_ (where values smaller than 1 indicate a decay).

### Country-wide, state-specific, and spatial analysis of average infection rates

To calculate and plot the overall (smooth) development of infection rates with time (c.f., Fig. 2 h) and depending on the weekday (Fig. 2 i), an appropriate GAMM (structurally strongly related to the above mentioned approach) has been applied. However, data of all ADs have been analysed simultaneously here, using again the federal state as a random intercept, but also the ADs (which are nested within the federal states). Furthermore, we introduced the date again as a smooth term, and the day of the week as a cyclic smooth (c.f., above). Model validation has been performed based on various residual plots (as e.g. suggested in [9, 19–21]).

**Figure 2.**
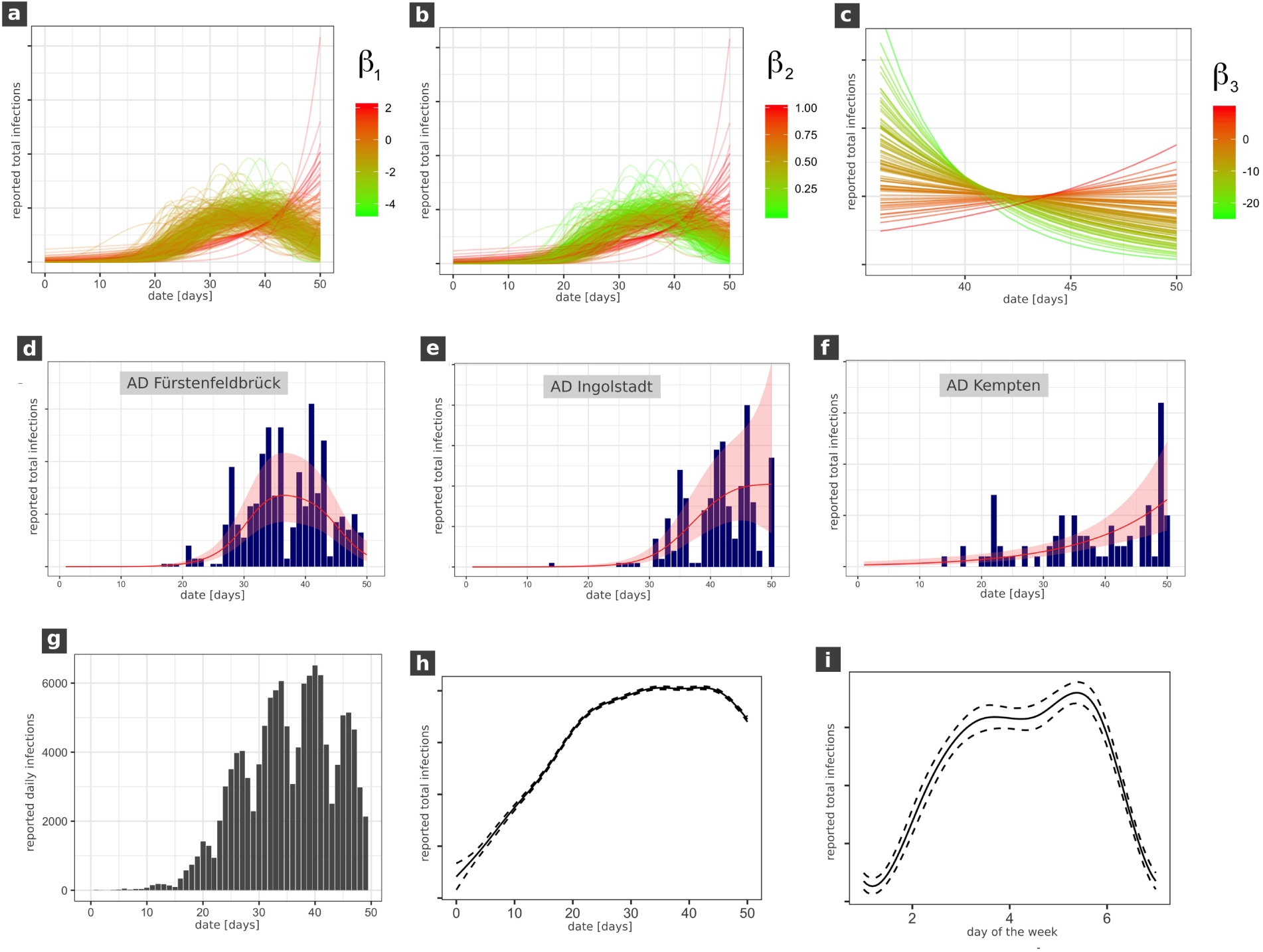
Temporal development of COVID-19 infection rates in Germany.(a)-(c) temporal infection curves separately estimated with a GAMM for each administrative district (AD). (a)-(b) total time series, (c) the last 14 days. The colour represents the corresponding *β*-value, where green colour indicates a recent decay and red colour a recent increase in infections. (d)-(f) example plots of reported infection numbers for three AD’s with a recent decay (d), saturation (e) and increase (f). (g) country-wide total daily reported infection numbers, (h)-(i) smoothed GAMM plot of total infection numbers, where (h) is the partial effect of the date, and (i) is the partial effect of the weekday. In all sub-figures (except (g)) the y axis cannot be interpreted in absolute values since either curves have been normalised for each AD, or partial effect plots are shown.

To analyse if the very recent decay of infection in Germany is significant, we used the three *β*-values as the outcome variable in three different appropriate GAMMs. In particular, we used a 2D thin spline regression depending on geographical coordinates to reduce spatial autocorrelation [18], and introduced the federal state as a random intercept. In particular, for *β*_1_ and *β*_3_ we used the Gaussian probability distribution, whereas for *β*_2_ we applied a Beta-distribution since this value is restricted between zero and one [3, 9]. Furthermore, for *β*_1_ and *β*_2_, data from different ADs have been weighted (using *a priori* regression weights) with respect to the total number of cases in the corresponding AD, whereas for *β*_3_ the inverse variance of the estimated regression coefficient has been used as a weight (‘inverse variance weighting’ [6]). Furthermore, the federal state has been used as a random intercept in all three models.

To analyse the average *β*-values for the different states (Fig. 3), in the above mentioned GLMMs (using the normal-respectively the Beta-distribution) the federal state has not been used as a random intercept but as a fixed effect predictor, using the federal state Baden-Württemberg as the baseline level. Furthermore, we neglected the spatial 2D smooth in order to avoid collinearities between the spatial smooth and the federal states. If smooth spatial maps of the *β*-values have been generated (Fig. 4), similar models have been used but now neglecting the federal state variable and using only a spatial 2D smooth as a predictor instead.

**Figure 3.**
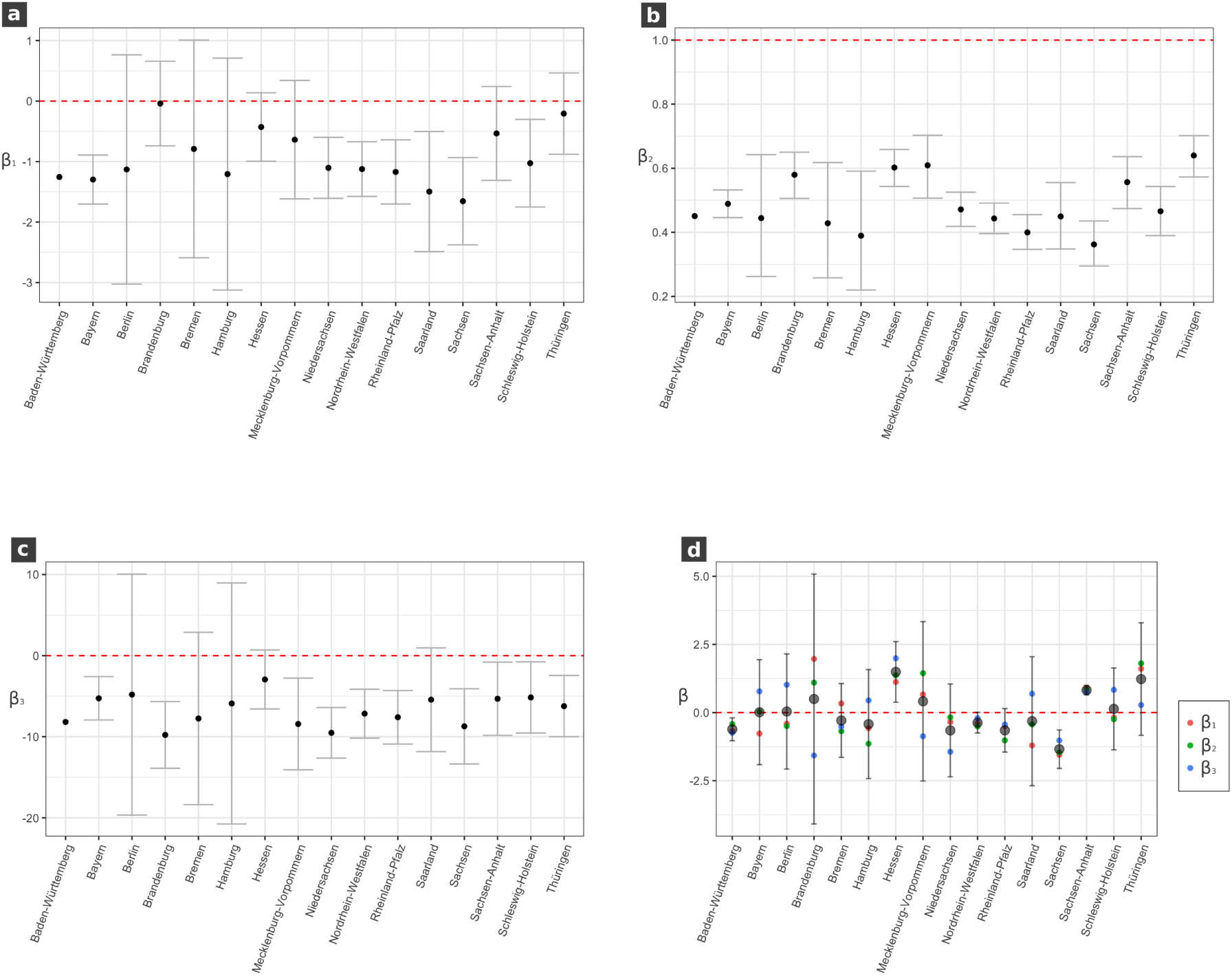
The GAMM-based estimated values of *β*_1_, *β*_2_ and *β*_3_ separately for each state. (a)-(c) separate estimates for each *β*. (d) mean values (grey large dots) and single values (coloured dots) of rescaled *β*-values for each state. Whiskers depict 95 %-confidence intervals (CIs). In (a)-(c), CIs are related to estimated *β*-values, in (d) they refer to the certainty of the average from all three rescaled *β*-values. For the state Baden-Würtetemberg in (a)-(c) no confidence intervals have been computes, since this state served as the baseline-level during GAMM-analysis.

**Figure 4.**
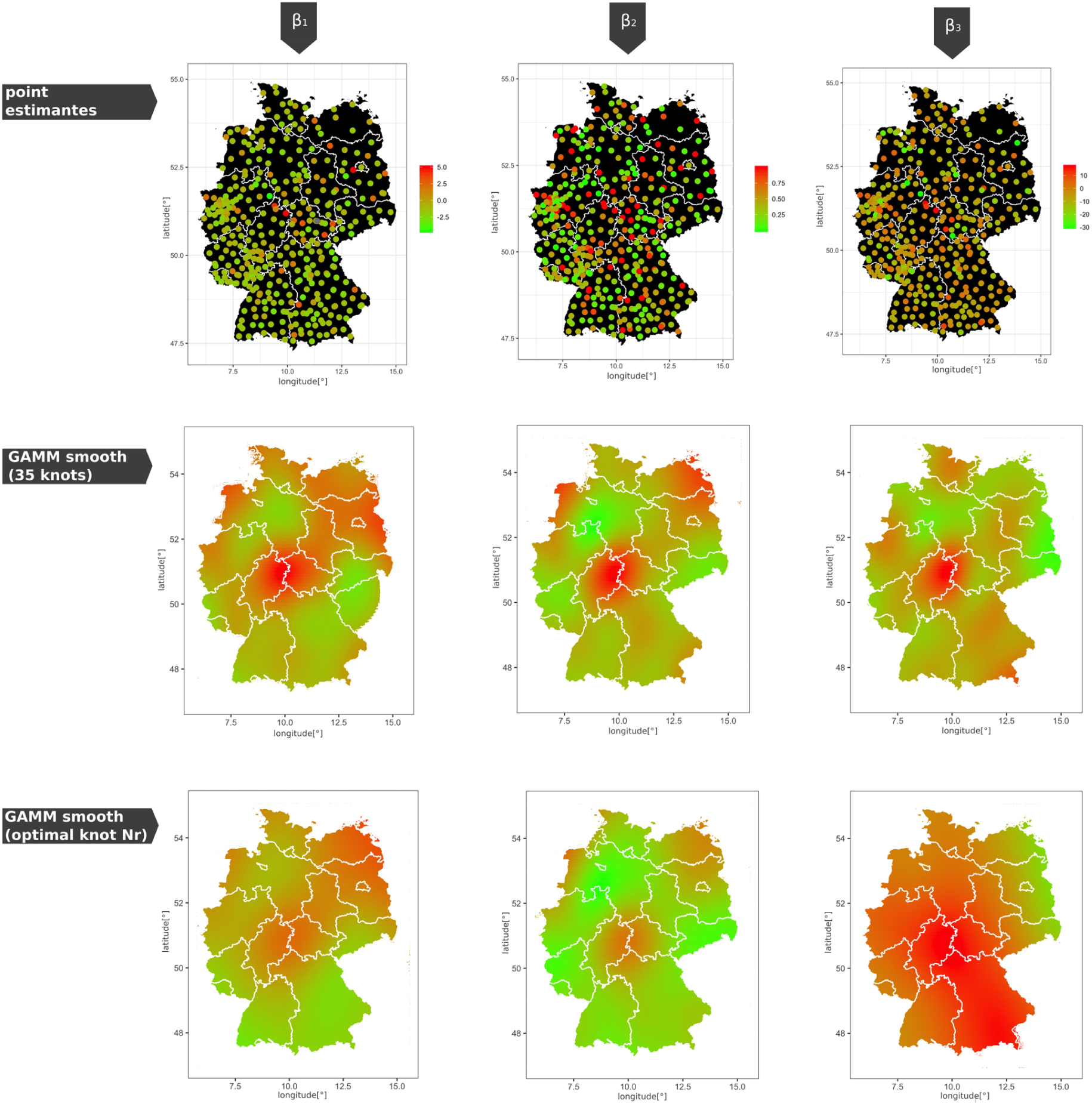
Spatial distribution of the three *β*-values. First row: raw data, second row: GAMM-smooth (where the number *k* of spatial knots has been forced to be *k* = 35), third row: GAMM-smooth (where the optimal number *k* has been estimated via generalised cross-validation).

### Selecting promising factors locally driving infection rates

In order to evaluate possible factors locally driving infection rates, we applied the ‘least absolute shrinkage and selection operator’ (LASSO) method [14, 15] in combination with cross-validation to the data. LASSO is known to reliably perform model selection even if the total number of possible predictors is high [9, 16]. In particular, we applied LASSO separately to three different types of models (using *β*_1_, *β*_2_, respectively *β*_3_ as the outcome variable, where for *β*_1_ and *β*_2_ we used the normal probability distribution. Since the used software (the R-package *glmnet*, c.f. below) is not able to perform Beta-regression, we reformulated for the LASSO-step the value *β*_2_ into a binomial variable instead (‘smaller one’ vs. ‘one’), using a binomial error distribution. We tested the following variables: On the AD-level we introduced *Longitude, Latitude* (as well as their interaction term), the date of the first reported COVID-19 infection (*first infection*), the local percentage of people older than 65 years (*Age*) (data from https://www-genesis.destatis.de), the population density (*Density*), and the local infection intensity (percentage of people infected by COVID-19 – *infection density*). On the coarser level of states, we introduced human mobility data. In particular, we used mobility data from Google collected by the type of activity, where activity (visits and length of stay) has been quantified at different types of places such as retail and recreation, groceries and pharmacies, parks, transit stations, workplaces, and residential – compared to a baseline level before the COVID-19 pandemic (variables have been named accordingly). The baseline has been calculated from a five-week period Jan. 3th until Feb. 6th, 2020. For the pandemic period, we averaged the values from Feb. 7th until the date of publication. Data have been obtained from https://github.com/datasciencecampus/google-mobility-reports-data/blob/master/csvs/international_local_area_trends_G20_20200410.csv.

### Software

For all statistical analyses we used the open source software R [12]. In particular, for GAMM regression analyses we used the functions *gam*() and *gamm*() from the R-package *mgcv* [18]. For LASSO analyses we used the function *glmnet*() from the R-package *glmnet* [5], and for all visualizstions we used the R-package ggplot2 [17].

### Infection rates significantly decrease in Germany

In Fig. 2 (a)-(c) the smoothed temporal development of COVID-19 infections separately estimated for each AD are plotted, where the colour represents the corresponding *β*-value representing the strength of decay (green) respectively increase (red) of infections. As described within the material and methods section, *β*_1_ and *β*_2_ are calculated based on the entire time frame (starting with February 22th), whereas *β*_3_ represents the percentage change in infection rates per day during the last 20 days. Three examples for differently developing sub-regions (ADs) are shown in (d)-(e) (all from the state Bayern): (d) shows a recent decay, (e) a saturation and (f) a recent increase. However, it is obvious that the majority of curves in (a)-(c) show a recent decay in infection rates. This recently decaying behaviour can also be seen when looking at the raw data of total (country-wide) infection numbers (2 (g)). Indeed, when estimating the average country-wide infection rate curve without the effect of the weekday (2 (h)) the rate decays since approximately 10 days, after being rather constant for 10-20 days before the decay. The partial effect of the weekday shows a distinct minimum at the weekend (2 (e)). New infections are first reported to the local health office and then collected by the RKI. During weekends, notifications of new infections are delayed and thus influence the RKI infection dataset, creating the observed decrease at weekends.

In order to analyse if the country-averaged decay is significant, we applied three appropriate GAMMs using each *β*-value as the outcome variable. Indeed, all three results indicate a recent highly significant country-wide decay: *β*_1_ = −1.00 (95 % CI: [−1.26, −0.74]), *β*_2_ = 0.50 (95 % CI: [0.45, 0.55]), and *β*_3_ = −6.76 (95 % CI: [−7.9, −5.7]). Just a reminder: for *β*_1_ and *β*_2_, negative values indicate a recent decay, where for *β*_2_, values smaller than one are associated with decreasing infection rates.

### Recent infection rates show a distinct spatial variability

Although decay has recently been observed in the country-wide infection rates, Fig. 2 (a)-(f) suggests that there is a strong local variability in that decrease. To analyse the relative differences between different states, we again used appropriate GAMMs; corresponding results are presented in 3 (a)-(c). In a second step, we rescaled each *β*-value across all states (i.e., subtracting the mean and dividing by the standard deviation) to make the relative differences between states comparable with respect to *β*_1_, *β*_2_ and *β*_3_. Corresponding results are given in 3 (d); the state-specific mean values from the rescaled *β*_1_, *β*_2_ and *β*_3_ thus represent the relative differences of recent infection rate changes between the different states. In particular, low values represent a strong decrease in infection rates, whereas high values represent a less strong decrease (respectively increase). The absolute values of the Y-axis in 3 (d) cannot be interpreted directly. Furthermore, confidence intervals in 3 (d) refer to the certainty of the mean value from the rescaled *β*_1_, *β*_2_ and *β*_3_ for each state.

It appears, that the states Sachsen and Bayern show the strongest (and most certain) decays in infection rates, whereas Thüringen and Hessen show the less-strongest decays. That Thüringen and Hessen nevertheless most probably show in average a decrease and not an increase, can be seen in 3 (a-c) where all estimated *β*_1_ and *β*_3_ values are below zero, and all *β*_2_ values are below one.

Spatial distribution of the different *β*-values is shown in Fig. 4. In the first row, the spatial raw data of the estimated *β*-values are shown. Sub-figures in the second row are based on spatial GAMM-plots, where the number of knots (= degrees of freedom) for the thin-plate regression spline has been fixed to *k* = 35 in all three plots. The aim is to obtain optimal comparability among the spatial distributions of *β*_1_, *β*_2_, and *β*_3_. In the third row, we present the optimal number of knots estimated separately for each *β*-value based on generalised cross-validation.

Overall, there is a very coherent picture of the spatial distribution of the different *β*-values: In all cases, the distribution is spatially inhomogeneous, showing in particular in the centre of Germany (on the boarder of Türinghen, and Hessen) a lower decay in infection rates compared to the average development. Indeed, both states show distinct larger beta values also in our analysis on the state-level (3). Only the cross-validation based GAMM-plot for *β*_3_ (4 third row, plot on the right-hand side) shows a much coarser spatial resolution. A possible explanation is that for the calculation of *β*_3_ much less data (namely infection data from only the last two weeks) have been used, which may lead to a larger unexplained variance, which finally leads to a more conservative estimate of spatial heterogeneity during the cross-validation procedure.

Thus, in summary, all states show in average a distinct decay in infection rates, where Sachsen and Bayern belong to the states with an especially strong decay, whereas in Thüringen and Hessen the decay-strength is less pronounced.

### Which factors drive the local decrease of infection numbers?

In order to further investigate the underlying factors causing the observed spatial differences in infection rates based on the proposed method, we tested different possible variables (c.f. Material and Methods section) with the LASSO variable selection method. However, the selection of variables was strongly determined by their public availability in an appropriate format, such that various potentially interesting factors have not yet been tested due to the lack of availability.

With respect to *β*_1_, this approach selected the variables *Infection density* and *Retail recreation*. The subsequent GAM-analysis with these predictors reveals a negative regression coefficient for *infection density* (-74.6, 95 %-CI: [-177.8,17.5]) suggesting that higher local infection densities lead to stronger current decays of infection rates. The psychological effect of perceived abstractness of the threat might be the reason for this: as long as the total number of infected in an area is low, it is highly probable that an individual does not know anyone infected personally and the threat is perceived to be very abstract. If in contrast an area is affected by a high number of infections, it his more probable that someone knows somebody who is infected (and/or the region is more present in the media) and the threat is suddenly perceived as very real, which may induce a much stronger change to a more preventive behaviour. Furthermore, there is a highly significant positive regression coefficient for *Retail recreation* (0.061, 95 %-CI: [0.02,0.1]) indicating that a high use of places like restaurants, cafes, shopping centres, theme parks, museums, libraries, and movie theatres prevents the recent decay of infection rates. Even when most of these places are closed now, a previous intense use of these places might still influence current rates due to the delay processes in infections.

With respect to *β*_2_, the LASSO analysis selected the variables *first infection, Age, infection density*, and *Transit stations*. The subsequent GAM-analysis with these predictors reveals highly significant dependencies on *infection density* and *first infection*. In particular, the positive regression coefficient for *first infection* (0.02, 95 %-CI: [0.01,0.03]) indicates that infection rates decay the stronger the earlier the first infection in this region has been detected. At first glance, one might think that an earlier start leads to an earlier decay due to the natural progression of the infection curve. However, the ‘natural progression’ would be a further increase for a long time, which only saturates or decreases to changes in behaviour. The latter can be caused either by intrinsic (psychological) or extrinsic factors (sanctions). Since sanctions (such as curfews) have been realised approximately at the same time everywhere in Germany (with only differences of some days), the observed decay in connection with early detected infections is also most probably a result of the above mentioned psychological factors.

A highly significant negative value for *infection density* (-97.2, 95 %-CI: [-135.6,-58.9]) suggest again that high local infection densities lead to stronger current decays of infection rates. Thus, both effects are most probably related to the above described awareness of the thread.

Furthermore, the positive regression coefficient for *Age* (0.022, 95 %-CI: [-0.002,0.045]) indicates that in regions with more people older than 65, infection rates decay less. This might be explained by the fact that younger people may show a more mobile behaviour (e.g. due to their work) so that sanctions (such as curfew) might affect them in a stronger way. Other factors might be that older people are possibly less frighten (due to more life experience), and it also could be more difficult for them to substantially change their habits.

The negative value for *Transit stations* (-0.012, 95 %-CI: [-0.027,0.002]) finally indicates that an increased use of public transport leads to a decrease of recent infection rates. This result is counter-intuitive, and we therefore think that here, possibly a random correlation was detected. Indeed, this result is statistically non-significant and movement data have been introduced only at the coarse federal state-level. For future work, such data with a finer spatial resolution (such as currently available for the UK https://www.gstatic.com/covid19/mobility/2020-04-05_GB_Mobility_Report_en.pdf) would allow for more reliably estimates.

Finally, with respect to *β*_3_, LASSO analysis did not select any variables. This observation might be again explained by the fact that for the calculation of *β*_3_ (compared to *β*_1_ and *β*_2_) much less data have been used (namely infection data from only the last two weeks instead of the entire period), which may lead to a larger amount of unexplained variance negatively influencing the detection of correlations during LASSO analysis.

## Summary and outlook

In summary, we present a new statistical approach which is specifically designed to identify factors influencing local infection rates. Since only relative changes in detected infections are considered, the approach is robust with respect to the percentage of undetected infections, as long as the latter does not locally change in time. The approach has been applied to spatio-temporal COVID-19 infection data in Germany. Nevertheless, it can be adapted to other countries at any state of infection, provided that both, infection numbers and factors affecting them, are provided in a sufficient spatio-temporal resolution. The analysis of daily COVID-19 infections in Germany reveals that (1) currently, infections are in average significantly decreasing; (2) there is a strong spatial variability in the local recent infection rates, especially varying in the centre of Germany (mostly including the two states Thüringen and Hessen); (3) there are strong evidences that both, an early emergence of first infections and high local infection densities, lead to strong recent decays in infection rates. The latter may indicate that psychological effects (such as the awareness of danger due to infection cases in the circle of friends and acquaintances) lead to a more cautious behaviour, which again reduces the local risk of additional infections.

The presented approach can be further applied in the nearest future to investigate additional potential variables, such as the impact of local restrictions (due to political decisions) or average movement range (based on cell-phone GPS data) on different spatio-temporal scales. For this study, the availability of such data was a bottleneck, e.g., since movement data are (up to our knowledge) not yet available on the spatial scale of administrative districts in Germany.

## Data Availability

All data evaluated in this study are publicly available

## Acknowledgments

The authors greatly thank Prof. Anna Marciniak-Czochra for revising the manuscript and fruitful discussions.

## References

1. S. Bhattacharya, M. M. Islam, and A. De. Search for trends of COVID-19 infection in India, China, Denmark, Brazil, France. Germany and the USA on the basis of power law scaling. medRxiv, 2020.

2. B. M. Bolker, M. E. Brooks, C. J. Clark, S. W. Geange, J. R. Poulsen, M. H. H. Stevens, and J.- S. S. White. Generalized linear mixed models: a practical guide for ecology and evolution. Trends Ecol Evol, 24(3):127–135, Mar 2009.

3. F. Cribari-Neto and A. Zeileis. Beta regression in R. 2009.

4. J. R. Donsimoni, R. Glawion, B. Plachter, and K. Wälde. Projecting the spread of COVID19 for Germany. 2020.

5. J. Friedman, T. Hastie, and R. Tibshirani. Regularization paths for generalized linear models via coordinate descent. Journal of statistical software, 33:1–22, 2010.

6. J. Hartung, G. Knapp, and B. K. Sinha. Statistical meta-analysis with applications. John Wiley & Sons. ISBN 978-0-470-29089-7., 2008.

7. T. Hastie and R. Tibshirani. Generalized Additive Models. London, UK: Chapman and Hall, 1990.

8. M. Kochanczyk, F. Grabowski, and T. Lipniacki. Dynamics of COVID-19 pandemic at constant and time-dependent contact rates. medRxiv, 2020.

9. F. Korner-Nievergelt, T. Roth, S. von Felten, J. Guelat, B. Almasi, and P. Korner-Nievergelt. Bayesian Data Analysis in Ecology Using Linear Models with R, BUGS, and Stan. Elsevier, London, 2015.

10. B. F. Maier and D. Brockmann. Effective containment explains sub-exponential growth in confirmed cases of recent COVID-19 outbreak in mainland China. arXiv: Populations and Evolution, 2020.

11. L. Peng, W. Yang, D. Zhang, C. Zhuge, and L. Hong. Epidemic analysis of COVID-19 in China by dynamical modeling. arXiv: Populations and Evolution, 2020.

12. R Core Team. R: A language and environment for statistical computing. R Foundation for Statistical Computing, Vienna, Austria., ISBN 3-900051-07-0, URL http://www.R-project.org/, 2020.

13. R. Schlickeiser and F. Schlickeiser. A Gaussian model for the time development of the SARS-COV-2 corona pandemic disease. Predictions for Germany made on march 30, 2020. medRxiv, 2020.

14. R. Tibshirani. Regression shrinkage and selection via the LASSO. J Royal Statisti Soci B, 58:267–288, 1996.

15. R. Tibshirani. The LASSO method for variable selection in the cox model. Stat Med, 16(4):385–395, Feb 1997.

16. M. Usai, M. Goddard, and B. Hayes. LASSO with cross-validation for genomic selection. Genet Res (Camb)., 91(6):427–36, 2009.

17. H. Wickham. ggplot2: Elegant Graphics for Data Analysis. Springer Dordrecht Hiedelberg London New York, 2009.

18. S. Wood. Generalized Additive Models: An Introduction with R. Chapman & Hall/CRC., 2006.

19. A. Zuur and E. Ieno. A protocol for conducting and presenting results of regression-type analyses. methods ecol evol 7: 636–645, 2016.

20. A. Zuur, E. Ieno, and G. Smith. Analysing Ecological Data. Springer Science+Business Media, LLC, 2007.

21. A. Zuur, E. Ieno, N. Walker, A. Saveliev, and G. Smith. Mixed Effect Models and Extensions in Ecology with R. Springer Science+Business Media, LLC, New York, 2009.

